# An early activated CD38^high^CD209^high^CD36^low^ monocyte phenotype is associated with progression to severe dengue

**DOI:** 10.64898/2026.11.06.26367260

**Authors:** Madushika Dissanayake, Malithi De Silva, Janarthani Arulnathan, Farha Bary, Thinuri Mithara Hapuarachchi, Ayeshika Lakmani Perera, Harshani Chathurangika, Chathura Ranatunga, Damayanthi Idampitiya, Thisara Perera, Eranga Narangoda, Ananda Wijewickrama, Chandima Jeewandara, Graham Ogg, Gathsaurie Neelika Malavige

**Affiliations:** Institute of Allergology and Immunology; Department of Immunology and Molecular Medicine, University of Sri Jayewardenepura, Gangodawila, Nugegoda, Sri Lanka; National Institute of Infectious Diseases, Angoda, Sri Lanka; MRC Translational Immune Discovery Unit, MRC Weatherall Institute of Molecular Medicine, University of Oxford, Oxford, United Kingdom

**Keywords:** dengue, monocytes, CD38, CD36, lipid uptake, obesity

## Abstract

**Background:** Although monocytes are among the principal target cells of dengue virus (DENV) infection we sought to investigate monocyte phenotypic markers associated with disease severity and evaluate their potential as prognostic biomarkers and therapeutic targets.

**Methods:** Differences in monocyte subpopulations and phenotypic markers were assessed by flow cytometry in patients with dengue fever (DF=33) and dengue haemorrhagic fever (DHF=21) and compared to health controls (HCs=20). Further in vitro experiments were done by co-culturing peripheral blood mononucleocytes (PBMCs) of healthy individuals (n=8) with DENV2, DENV3, NS1 antigen of both serotypes and LPS. The changes in expression of CD36 and CD38 in vitro in monocytes were assessed.

**Results:** A significant expansion of intermediate monocytes and a contraction of the classical monocytes was seen in patients with DHF compared to DF and HCs, with no significant differences within the non-classical monocytes. Patients who progressed to develop DHF, exhibited an activated monocyte phenotype characterized by, expansion of CD38^high^, CD209^high^, CD86^high^, HLA-DR^high^, CD36^low^, intermediate monocyte subset. The highest expression of CD38 and CD86 was seen in non-classical monocytes, and CD209 and HLA-DR in intermediate monocytes. Most significant downregulation of CD36 was observed in non-classical monocytes. CD38 expression on monocytes (especially classical monocytes) positively and significantly correlated with liver transaminases and serum ferritin levels. Co-culture of PBMCs with DENV3 led to significant downregulation of CD36 (p=0.023).

**Conclusions:** These results suggest that early alterations in monocyte activation, antigen presentation, viral entry, and lipid sensing pathways contribute to dengue immunopathogenesis.

## Introduction

Dengue is the fastest spreading mosquito borne viral infection worldwide, with an estimated 5.66 billion living in areas suitable for dengue and other arbovirus transmission [1]. Due to climate change resulting in higher global temperatures and rapid and unplanned urbanization, the dengue incidence has markedly increased in endemic countries and has spread to new geographic locations [2]. In parallel, the age-stratified dengue incidence, mortality rates and disability adjusted life years have risen, especially in South Asia, Southeast Asia and Latin America[3]. Although most individuals infected with the dengue virus (DENV) develop an inapparent dengue infection, 25-30% of individuals develop symptomatic disease, which can lead to develop complications such as dengue haemorrhagic fever (DHF), shock, bleeding and organ dysfunction, contributing to fatalities [4]. While timely of plasma leakage and meticulous, individualized fluid management have dramatically reduced case fatality rates, mortality remains higher during large outbreaks owing to the overwhelming demand on healthcare services. As there are no biomarkers to predict those who will develop plasma leakage and no specific treatment for dengue, it would be crucial to find both prognostic markers and therapeutic targets to reduce hospitalizations and progression to severe disease.

Following inoculation of the DENV into the skin by the Aedes mosquitoes, the virus initially infects immune cells within the dermis where virus replication facilitates dissemination into the circulation and development of viraemia [4]. Monocytes are among the principal circulating target cells [5, 6]. Monocytes are directly infected by the DENV either through binding to CD209 (DC-SIGN), or internalized through Fcγ receptors, when bound to poorly neutralizing DENV-specific antibodies, resulting in antibody dependent enhancement (ADE) [7, 8]. In addition, monocytes can also be infected through TLR2 dependent mechanisms, which have shown to associate with clinical disease severity [9]. Once infected, monocytes produce a vast array of inflammatory cytokines and chemokines, which are associated with endothelial dysfunction, and inflammation [10, 11]. Monocytes can produce vast amounts of IL-10 which has shown to associate with severe disease and T cell dysfunction[12, 13], when infected through ADE [14]. These findings indicate that monocytes are key contributors to dengue pathogenesis, making them attractive targets for therapeutic intervention.

There are many studies investigating the role of monocytes in acute dengue. While some studies have characterized the changes in the different monocyte subsets, showing a reduction of classical monocytes (CD14^high^ CD16^low^) and non-classical monocytes (CD14^low^CD16^high^) with an expansion in intermediate monocytes (CD14^high^CD16^high^) [15–17], some have characterized monocytes in greater detail by carrying out single-cell RNA sequencing studies [5, 18]. Single-cell RNA sequencing studies of patients with acute dengue have shown that classical and intermediate monocytes upregulate genes involved in endothelial dysfunction, chemotaxis, angiogenesis, and leukocyte extravasation, suggesting that these cells contribute to dengue pathogenesis [18]. Other studies which have investigated expression of activation markers, co-stimulator receptors and antigen presentation molecules (including HLA-DR, CD86, CD80, CD32, CD64) have shown varying results, possibly due to differences in the day since onset of illness at patient recruitment and differences in disease severity [15, 19].

Although monocytes are recognized as key mediators of dengue pathogenesis, the expression and clinical significance of many emerging monocyte phenotypic markers remain largely unexplored in acute dengue. Investigating these markers may improve our understanding of disease mechanisms and identify new targets for therapeutic intervention. Therefore, in a cohort of Sri Lankan patients with varying severity of acute dengue, we investigated the differences in the phenotype of monocytes, and expression of previously unexplored markers such as CD38, CD36 and CD209 expression and further carried out in vitro experiments to understand if the DENV could lead to altered expression of these markers on monocytes.

## Methods

### Recruitment of patients

54 adult patients with acute dengue, who presented within ≤5 days since onset of illness, were recruited following informed written consent from the National Institute of Infectious Diseases, Sri Lanka, between August 2024 and June 2025. The day of onset of illness was defined as the day of onset of fever. Patients with known chronic kidney or liver disease were excluded. None of the patients had evidence of plasma leakage at the time of recruitment. Patients were followed up from the day of admission to discharge and all clinical features (presence of fever, dengue warning signs, blood pressure, pulse rate, urine output) and laboratory characteristics (full blood count), were measured several times a day from the day of admission to discharge. As practiced in this hospital, all dengue patients were subjected to ultrasound scans of the abdomen daily to detect presence of fluid leakage. A diagnosis of DHF was made based on the presence of fluid leakage during the hospital stay, while those who did not develop fluid leakage and did not fulfill the criteria of DHF were classified as having DF based on the WHO dengue disease classification [20].

Accordingly, 21/54 were classified as having DHF, and 33 were classified as having DF. To determine the differences in monocyte subsets and phenotypic markers, 20 healthy adult volunteers were recruited from the University of Sri Jayewardenepura staff following written informed consent. Dengue was confirmed by a positive NS1 antigen test (SD Biosensor, South Korea) a positive result by DENV-specific real-time PCR.

### Anthropometric measurements

Anthropometric measurements were obtained from all patients at the time of recruitment. The weight was recorded using a digital scale with participants barefoot and in light clothing. Height was measured barefoot with a stadiometer, and waist circumference was obtained using a non-stretchable tape. Those with a BMI ≥23.9 kg/m² were defined as being overweight and those with a waist circumference ≥80 cm for women or ≥90 cm for men were classified as having central obesity, based on the WHO classification of obesity and central obesity in South Asian populations [21].

### Quantification of IL-18 and ferratin levels

IL-18 and ferritin were measured in serum samples stored at -80°C. IL-18 ELISA was performed according to the manufacturer’s instructions (R&D Systems, Bio-Techne, Minneapolis, MN, USA). A four-parameter logistic regression model was used to interpret the assay results for the IL-18 assays. Serum ferritin levels were quantified using the AutoLumo A1860 analyzer (AutoBio, China), a chemiluminescence immunoassay-based automated system. The assay was conducted according to the manufacturer’s protocol, ensuring high specificity and sensitivity for ferritin detection. Calibration and quality control procedures were performed prior to analysis to maintain assay accuracy.

### Phenotypic analysis of monocytes using flow cytometry in patients with acute dengue

To assess the differences in monocyte subsets in relation to disease severity and obesity, a multicolor flow cytometry panel was designed to provide a comprehensive profile of monocyte heterogeneity and immune responsiveness. CD14 (FITC, BioLegend, USA) and CD16 (Pacific Blue, BioLegend, USA) were used to distinguish classical, intermediate, and non-classical monocytes, CD38 (Alexa Fluor700, BioLegend, USA), CD86 (PE, BioLegend, USA), and HLA-DR (APC, BioLegend, USA) to identify activation and antigen-presenting capacity and CD209/DC-Sign (PE Cy7, BioLegend, USA) and CD36 (APC Cy7, BioLegend, USA) were used to determine expression of one of the DENV receptors and lipid uptake functions. To exclude lineage positive cells, CD3, CD19, CD15, CD56 (BV 605, BioLegend, USA) were used and fixable viability dye Zombie Aqua (BV 510, BioLegend, USA) to exclude dead cells during the analysis. Antibody titrations were performed prior to staining to determine optimal concentrations. The clones and manufacture are shown in Supplementary methods table 1. The gating strategy for identification of the different markers are shown in supplementary figure S1. Gates for each marker were defined using the corresponding FMO (Fluorescence Minus One) control to distinguish true positives. FMO controls for specific markers are shown in supplementary figure S2.

Whole blood samples of patients and healthy individuals were used for phenotypic analysis of monocytes. Briefly, the samples were subjected to red blood cell lysis (using BD FACS lysing solution), and fresh cells were washed and stained for 30 minutes in ice-cold PBS containing 2% FBS with the above antibodies. Flow cytometry data was acquired using a BD FACSAria III cytometer (BD Biosciences) and analysed using FlowJo software (version 10).

### In vitro assays on monocytes to determine effect of the DENV and NS1 on CD36 and CD38 expression

Peripheral Mononuclear cells (PBMC) were isolated using density-gradient centrifugation method from whole blood of 8 healthy individuals. PBMC were co-cultured (1×10^6^ cells in media) with either DENV2 at and MOI of 0.5 and DENV3 at an MOI 0.1, DENV2 and DENV3 NS1 antigen at 2µg/ml (Native Antigen, UK), LPS (0.02 µg/ml, Invivogen, USA) or with media alone (control). The media contained RPMI-1640 (Capricorn Scientific, Ebsdorfergrund, Germany), 2 mM L-glutamine (Sigma-Aldrich, USA), 1% penicillin-streptomycin (Sigma-Aldrich, USA) and 10% FBS (Sigma-Aldrich, USA). The virus strain used, virus isolation and determining MOI is described in supplementary methods. The cells were incubated at 5% CO2 at 37^0^C and were harvested at 24 hours for analysis by flowcytometry. The cells were stained with anti-CD14-FITC (BioLegend, USA), anti-CD16-PacificBlue (BioLegend, USA), anti-CD36-APC-Cy7 (BioLegend, USA) and anti-CD38-AlexaFlour700 (BioLegend, USA) along with the fixable viability dye Zombie Aqua (BV 510, BioLegend, USA) to analyze the surface expression of CD36 and CD38 on monocytes. The results were analyzed in FlowJo version 10. The gating strategy used in the analysis is shown in supplementary figure S3.

### Statistical analysis and post-hoc analysis

Statistical analyses were performed using GraphPad Prism version 9.5.1 (Dotmatics, California, USA). Data distribution was assessed using the Shapiro–Wilk test, and because all variables were non-normally distributed, results are reported as medians with inter-quartile ranges (IQR). Comparisons between two independent groups were performed using the two-tailed Mann–Whitney U test, while comparisons involving more than two groups were conducted using the Kruskal–Wallis test followed by Dunn’s post-hoc multiple comparison test.

Associations between monocyte markers and clinical parameters (IL-18, ferritin, AST, ALT, and CRP) were assessed using non-parametric Spearman rank correlation. Correlation matrices were visualized as heat maps generated in R (version 4.3.2) using the heatmap package. A two-tailed P value < 0.05 was considered statistically significant. P-values were corrected for multiple comparisons using the Benjamini-Hochberg method.

High-dimensional data analysis including data preprocessing and transformation, generation of high-dimensional structure of data in two dimensions, including Uniform Manifold Approximation and Projection (UMAP) algorithm using biomeStat (Bioinformatics Analysis Platform. Version 1.0, Advance Molecular Works. Inc). Identification of clustering and identification of populations are described under supplementary methods.

CD36 and CD38 MFI were measured at 0 and 24 hours following *in vitro* stimulation of PBMCs with NS1, LPS, DENV2, or DENV3, alongside an unstimulated (Nil) control. For each stimulus, the change in MFI (24h − 0hours) was compared to the change under the Nil condition using the paired Wilcoxon signed-rank test, isolating the effect of each stimulus from non-specific changes occurring over time in culture. P-values were corrected for multiple comparisons across the three stimuli (NS1, LPS, DENV2/DENV3) within each marker using the Holm-Bonferroni method. This analysis was performed in Python (version 3.11.9).

## Results

### Patient characteristics

The patient characteristics of the 21 patients with DHF and 33 patients with DF are shown in supplementary results table 1. The mean age of patients with DF, DHF and healthy controls (HCs) was similar. 24 (44.4%) of patients were males. Both patients with DF and DHF were recruited on median day 4 (IQR, 3-5) since onset of illness to capture monocyte changes in early illness.

### Differences in monocyte sub-sets and phenotypic markers in patients with varying severity of acute dengue

Significant differences in the distribution of monocyte sub-populations were seen in patients with acute dengue compared to healthy individuals (n=20) and between patients with DF and DHF (Figure 1A, B). The classical monocytes were significantly contracted in patients with DHF (p<0.0001) and DF (p=0.01) compared to HCs, with also significant contraction in patients with DHF compared to DF (p=0.001). In contrast, the intermediate monocyte subpopulation was significantly expanded in patients with DHF (p<0.0001) and DF (p=0.0002) compared to HCs. Patients with DHF also had significantly higher intermediate monocytes than patients with DF (p=0.004).

**Figure 1:**
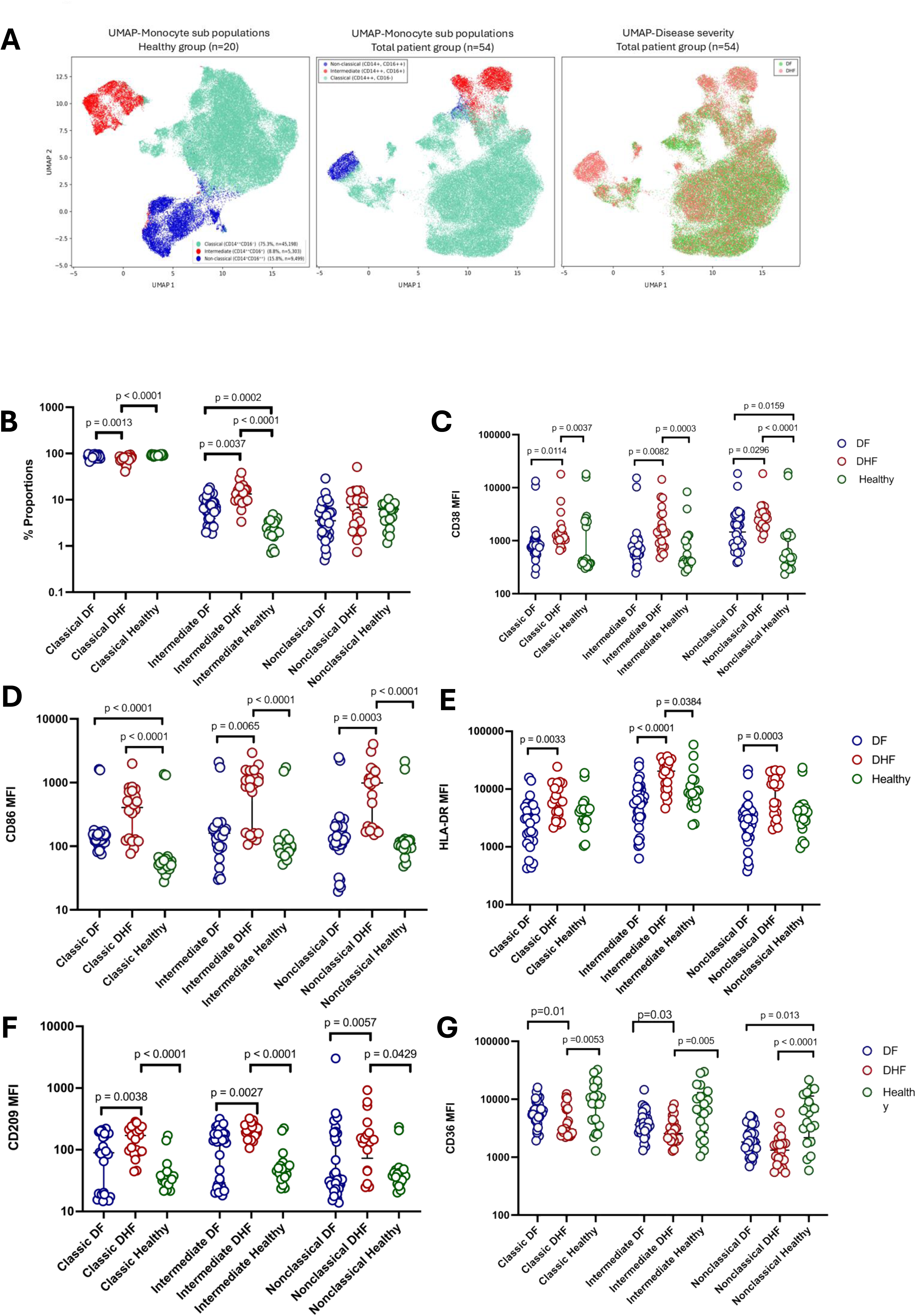
Phenotypic differences in different monocyte subpopulations in patients with varying severity of acute dengue. Patients with DF (n=33) and DHF (n=21), were recruited in acute illness and different monocyte subpopulations were compared with healthy individuals (n=20) by flowcytometry. The differences were visualized by using a Uniform Manifold Approximation and Projection (UMAP) algorithm (A), and the differences were also shown for patients with DHF, DF and healthy controls (B). The differences in expression of CD38 (C), CD86 (D), HLA-DR (E), CD209 (F) and CD36 (G) between different patient groups and monocyte subpopulations were determined. The statistical significance was determined by the Kruskal–Wallis test followed by Dunn’s post-hoc multiple comparison test.

CD38 is shown to be upregulated in activated monocytes and macrophages, and has multiple functions including binding to platelets, endothelium and increase in glycolytic activity and cytokine production by monocytes [22]. CD38 was seen to be significantly upregulated in all monocyte subpopulations in patients with DHF, compared patients with DF and HCs, with highest expression levels seen in non-classical monocytes (Figure 1C).

CD86 is a co-stimulatory molecule that interacts with CD28 and CTLA-4 on T cells and is upregulated upon activation of monocytes [23]. As seen with CD38, CD86 was significantly upregulated in all monocyte subpopulations in patients with DHF, compared to HCs, with the highest expression seen in non-classical monocytes (Figure 1D). Expression rates were significantly higher in patients with DHF compared to DF in non-classical and intermediate monocytes.

HLA-DR is an MHC-class II molecule involved in antigen presentation to CD4+ T cells and thereby plays a significant role in initiating T cell responses [24]. Although there were no differences in HLA-DR expression between patients with DF and HCs, HLA-DR expression was significantly upregulated in all monocyte subsets in patients with DHF, compared to those with DF (Figure 1E).

CD209 (DC-SIGN) is expressed on dendritic cells and monocytes and has been identified as an important receptor for entry of the DENV into these cells, through virus internalization [8] . CD209 expression was significantly upregulated in all monocyte populations in patients with DHF, compared to HCs (Figure 1F). Expression of CD209 was significantly higher in non-classical monocytes in patients with DHF compared to patients with DF and HCs. The highest expression levels were seen in intermediate monocytes in patients with DHF.

CD36 is a scavenger receptor expressed on monocytes and many other immune cells and binds to oxidized phospholipids including oxidized low density lipoprotein (oxLDL) [25]. Together with TLR4 and TLR6, CD36 has shown to activate NLRP3 inflammasome and thereby induce inflammation[25]. However, downregulation of CD36 is known to reduce the ability of monocytes to phagocytose apoptotic neutrophils[26]. We found that CD36 was downregulated in both patients with DF and DHF compared to HCs, with significant downregulation in patients with DHF (Figure 1G). Significant downregulation was seen in patients with DHF compared to DF. The lowest expression of CD36 was observed in non-classical monocytes in patients with DHF.

Collectively this data shows that patients who progressed to develop DHF, exhibited an activated monocyte phenotype characterized by, expansion of CD38^high^, CD209^high^, CD86^high^, HLA-DR^high^, CD36^low^, intermediate monocyte subset.

### Association of phenotypic changes in monocytes in patients with varying severity of acute dengue with inflammatory mediators

It has been shown that higher ferritin, C-reactive protein (CRP), liver transaminase levels were associated with increased clinical disease severity [27, 28]. We found that CD38 expression on classical monocytes was significantly and positively correlated with serum AST (Spearman’s R=0.53, adjusted p=0.001), ALT (Spearman’s R=0.42, adjusted p=0.036), and serum ferritin (Spearman’s R=0.47, adjusted p=0.026) levels (Figure 2). Although not significant, CD36 expression on different monocyte subsets, was negatively associated with ferratin and IL-18 and also with AST and ALT.

**Figure 2:**
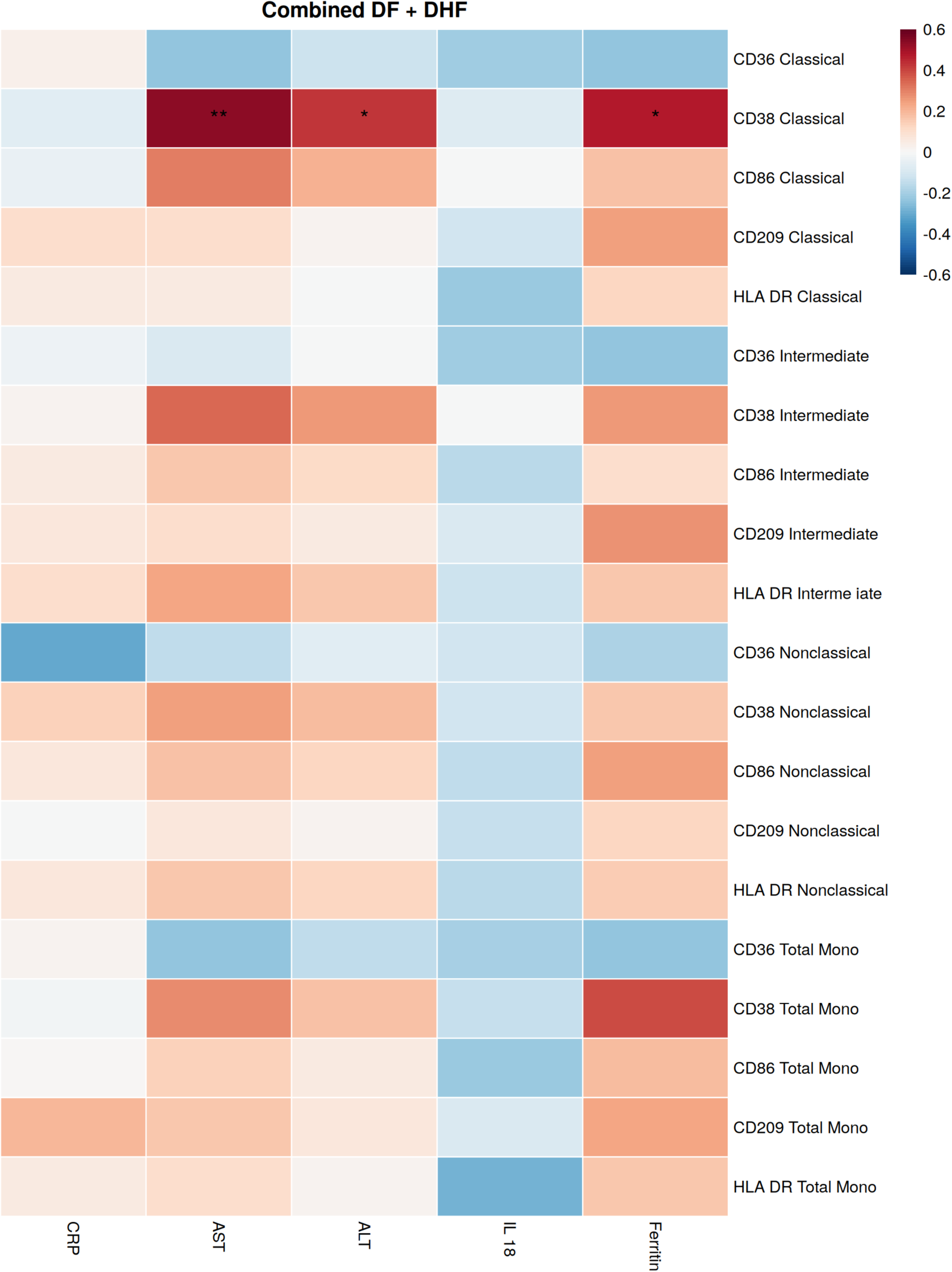
Association of monotypic phenotypic markers with inflammatory mediators and liver transaminases. Monocyte phenotypic markers were correlated with AST, ALT, serum ferratin, IL-18 and CRP levels. Correlation matrices were visualized as heat maps generated in R (version 4.3.2) using the heatmap package. A two-tailed P value < 0.05 was considered statistically significant.

### Differences in monocyte sub-sets and phenotypic markers in obese vs lean patients with acute dengue

We and others have previously found that obese patients are significantly more likely to develop severe dengue, associated with dysfunctional T cell and NK cell responses [27, 29, 30]. Therefore, we sought to determine if there were differences in monocyte subsets and phenotypic markers in obese (n=25) vs lean (n=28) patients with acute dengue. We did not observe any differences in monocyte subsets (Figure 3A) or expressions of CD86, HLA-DR, CD209 or CD36 in obese vs lean patients. However, lean patients had significantly higher expression of CD38 in intermediate monocytes than obese patients (Fig 3B).

**Figure 3:**
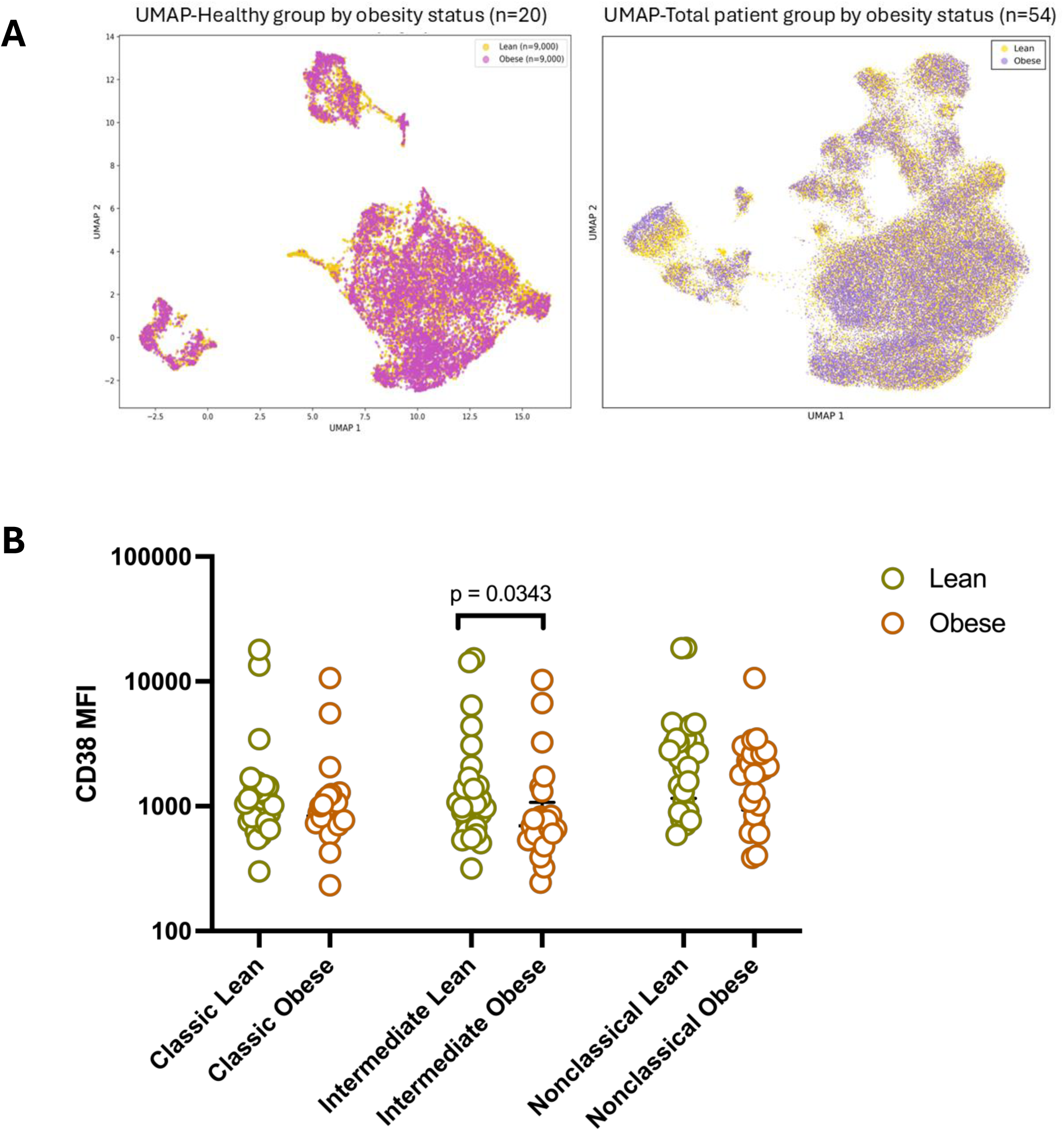
Phenotypic differences in different monocyte subpopulations in obese and lean patients. Differences of monocytes subpopulations were assessed by flowcytometry in patients with acute dengue who were obese (n=25) or lean (n=28). The differences were visualized by using a Uniform Manifold Approximation and Projection (UMAP) algorithm (A), and other phenotypic markers, where significant differences were seen for CD38 expression only (B). The statistical significance was determined by the Kruskal–Wallis test followed by Dunn’s post-hoc multiple comparison test.

### Effect of the DENV and NS1 on CD36 and CD38 expression on monocytes

As we observed significant downregulation of CD36 with upregulation of CD38, we sought to understand if these changes observed were due to infection with the DENV or due to interactions with the dengue NS1 protein. For these experiments we used both DENV2 and DENV3 serotypes, as it has been shown that recognition of the DENV2 by TLR2 is strain dependent [9] and engagement of TLR2 ligands lead to downregulation of CD36[26]. We found that while there was downregulation of CD36 with both DENV2 and DENV3 (Figure 4A, B), with significant downregulation of CD36 with DENV3. There was no significant downregulation of CD36 with either NS1 or LPS. Although in patients we observed CD38 upregulation, in vitro we saw a trend toward downregulation of CD38, although this was not significant.

**Figure 4:**
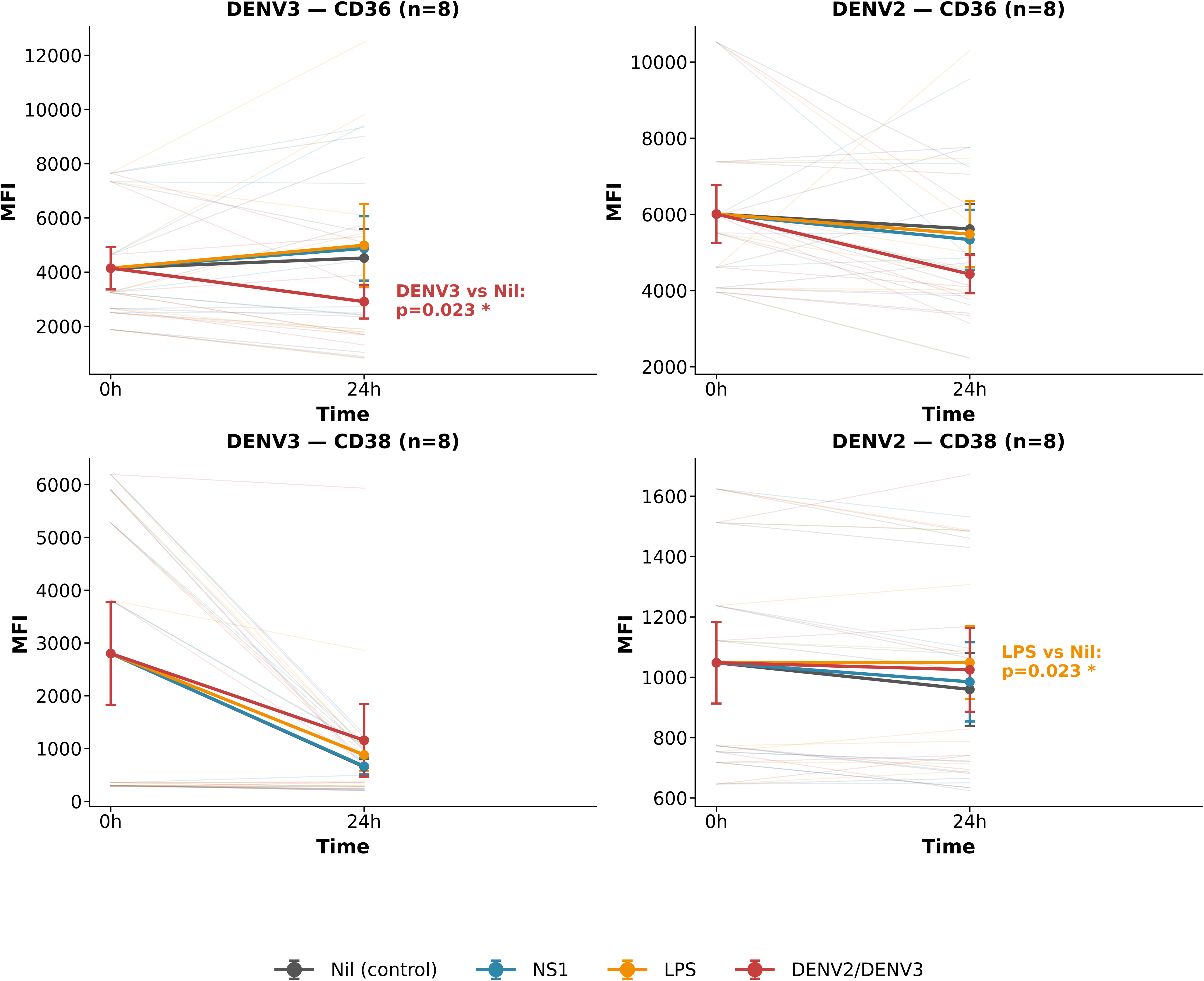
Differences in CD38 and CD36 expression in in vitro when co-cultured with DENV2 or DENV3, NS1 antigen of the 2 serotypes and LPS. PBMCs were co-cultured with either DENV2 or DENV3, with NS1 of both serotypes and with LPS for 24 hours and expression of CD36 and CD38 on monocytes assessed by flowcytometry. For each stimulus, the change in MFI (baseline to 24 hours) was compared to the change under the Nil (unstimulated) condition using the paired Wilcoxon signed-rank test, isolating the effect of each stimulus from non-specific changes occurring over time in culture. P-values were corrected for multiple comparisons across the three stimuli (NS1, LPS, DENV2/DENV3) within each marker using the Holm-Bonferroni method.

## Discussion

In this study we have examined certain novel phenotypic markers of monocytes in patients with acute dengue and identified markers that associate with clinical disease severity and varying metabolic status. We found that the intermediate monocyte population was significantly expanded in patients with acute dengue, with highest expansion in patients with DHF, along with lower proportion of classical monocytes as reported in other studies [16, 18, 31]. Therefore, DHF was associated with an expansion of the intermediate subset of monocytes displaying an activated monocyte phenotype characterized by an CD38^high^, CD209^high^, CD86^high^, HLA-DR^high^, CD36^low^ phenotype.

There are many studies describing the phenotype of monocytes in acute dengue [16, 18], including transcriptomic profiling of the different monocyte subpopulations to understand the functional characteristics of these subpopulations in relation to dengue disease severity [18]. These studies have shown that different monocyte subpopulations upregulate genes in relation to inflammation, interferon responses and chemotaxis to varying degrees [18] with both classical and intermediate monocytes upregulating genes related to endothelial dysfunction and leucocyte extravasation. To further understand the role of monocytes in dengue, as they have been shown to be one of the cell types that are predominantly infected by the DENV[5, 6], we studied known markers such as CD38, CD209 (DC-SIGN) and CD36 that had not been previously studied in relation to dengue disease severity to identify potential novel severity prediction and therapeutic targets. CD38 was found to be significantly upregulated in all monocyte subpopulations with the highest expression seen in patients with DHF. An increase in CD38^high^ monocytes was shown to be an early and an important marker of sepsis, which discriminated sepsis from sterile inflammation and it is also an important immunometabolism and calcium signalling. It has also been identified as a potential early biomarker for sepsis and increased expression was shown to associate with increased 28-day mortality. Although we found CD38 on monocytes in acute dengue was associated with disease severity, DENV *in vitro* was not associated with an upregulation of CD38. Therefore, CD38 upregulation in acute dengue could be due to different mechanisms, such higher levels of IFNγ and LPS found in patients with DHF [32, 33]. In lupus prone mouse models, it has been shown that inhibition of CD38 improved T cell function and viral clearance when infected with LCMV and improved mitochondrial function and defects in autophagy[34]. The DENV exploits the cellular autophagy pathways for viral replication, later leading to defects in autophagy [35, 36]. Furthermore, impaired T cell function which associated with delayed virus clearance has shown to associate with dengue disease severity[30, 37]. Therefore, it would be important to explore the possibility of utilization of CD38 as a prognostic marker for progression to severe dengue and also if it can be targeted by specific therapeutics in dengue.

CD36 is a class B scavenger receptor expressed on many immune cells and recognizes many damage associated molecular patterns, pathogen associated molecular patterns and also acts as a fatty acid transporter by binding to long chain fatty acids [25]. We found that CD36 was downregulated in all monocyte subpopulations with the lowest expression seen in patients with DHF. The DENV has shown to bind to TLR2 [9], while NS1 has shown to bind to TLR4[38]. Activation of TLR2 and TLR4 was shown to downregulate CD36 [26]. As we hypothesized that sensing of the DENV by TLR2 of monocytes could lead to downregulation of CD36 expression, we co-cultured both DENV2 and DENV3. These experiments did demonstrate that the virus led to downregulation of CD36. However, we did not investigate if the downregulation seen in vitro was due to the activation of the TLR2 signaling pathways. Nevertheless, the significant downregulation of CD36 in patients with acute dengue is likely to have consequences in monocyte function. For instance, monocytes which have downregulated CD36 have shown to be less efficient in phagocytosis of apoptotic neutrophils [26]. Furthermore, CD36 is an important pattern recognition receptor (PRR), and CD36 ^-/-^mice showed higher parasite loads, higher mortality and lower natural killer cell function than wild-type mice in malaria mouse models [39]. HIV-1 Nef protein has shown to downregulate CD36 in monocyte-derived macrophages, leading to impaired monocyte function[40]. These findings raise the possibility that DENV-mediated downregulation of CD36 contributes to impaired monocyte function.

DC-SIGN (CD209) is a receptor for the DENV and blocking this receptor has shown to significantly reduce virus infection of dendritic cells and other cells expressing this receptor [8]. Although DC-SIGN is an important PRR, many viruses such as the DENV hi-jack this to enter cells [41]. Polymorphisms of this receptor has shown to associate with disease severity[42]. Although the importance of this receptor in DENV infection has been well established, the differential expression of this receptor in relation to clinical disease severity had not been documented. DC-SIGN expression was significantly higher in patients with DHF when compared to both healthy controls and patients with DF. Upregulation of DC-SIGN in monocytes/dendritic cells has shown to associate with disease severity in patients with SARS-CoV-2 and influenza mouse-models [43, 44]. Senescent monocytes were shown to upregulate DC-SIGN expression by IL-10 induced JAK2-STAT-3 signaling pathways thereby becoming more susceptible to DENV infection[45]. We and other have shown that IL-10 is sone of the cytokines which were shown to significantly associate with disease severity[13, 46]. Given that dengue is predominantly now seen in adults with increased morbidity and mortality in older adults[47], increased DC-SIGN expression on monocytes and dendritic cells could be contributing to increased disease pathogenesis.

The other phenotypic markers CD86 and HLA-DR have been previously characterized by many groups. Our findings are similar to other groups which show that CD86, a co-stimulatory molecule involved in T cell activation, is expressed in higher frequencies in patients with DHF[17, 19]. Although high expression of CD86 reflects enhanced antigen presentation, thereby leading to more enhanced activation of antigen-specific T cells, many studies have shown that T cells in patients with DHF have impaired functionality [29, 30]. We observed that HLA-DR too was also significantly upregulated in patients with DHF compared to healthy individuals and patients with DF. Our findings are in contrast with the findings of Azeredo et al[48] and Naranjo-Gomez et al[15], due to differences in clinical disease severity classification and sampling time, from the onset of illness. As we recruited patients at earlier time points (≤5 days since onset of illness), it appears that upregulation both CD86 and HLA-DR indicating better antigen presenting capacity within antigen presenting cells. It would be important to longitudinally study these makers, in large cohorts and in children to further understand how monocyte function evolves over time in patients with varying severity of illness.

In summary, we show that acute dengue is associated with a distinct activated monocyte phenotype characterized by expansion of intermediate monocytes, upregulation of CD38, CD86, HLA-DR and CD209, and downregulation of CD36, with these changes being most pronounced in patients who develop DHF. These results suggest that early alterations in monocyte activation, antigen presentation, viral entry, and lipid sensing pathways contribute to dengue immunopathogenesis and identify several novel phenotypic markers that may serve as prognostic biomarkers or potential therapeutic targets for severe dengue.

## Availability of data and material

All data is available within the manuscript and the supporting files.

## Acknowledgements: Author contributions

Conceptualization and design: GNM, MD, GSO

Data curation: TMH, ALP,

Investigation: MD, JA, FB, CR

Resources: DI, TP, EN, CJ

Formal analysis: MD, MDS

Project administration: GNM, CJ, AW

Funding acquisition: GNM, CJ, GSO

Writing the initial draft: GNM, MD, MDS

Writing-reviewing and editing: GSO, GNM

## Data Availability

All data is available within the manuscript and the supporting files.

## References

1. Lim A, Shearer FM, Sewalk K, et al. The overlapping global distribution of dengue, chikungunya, Zika and yellow fever. Nat Commun 2025; 16:3418.

2. Malavige GN, Sjo P, Singh K, et al. Facing the escalating burden of dengue: Challenges and perspectives. PLOS Glob Public Health 2023; 3:e0002598.

3. Zhang WX, Zhao TY, Wang CC, et al. Assessing the global dengue burden: Incidence, mortality, and disability trends over three decades. PLoS neglected tropical diseases 2025; 19:e0012932.

4. Malavige GN, Ogg GS. Molecular mechanisms in the pathogenesis of dengue infections. Trends Mol Med 2024.

5. Ghita L, Yao Z, Xie Y, et al. Global and cell type-specific immunological hallmarks of severe dengue progression identified via a systems immunology approach. Nature immunology 2023; 24:2150–63.

6. Zanini F, Robinson ML, Croote D, et al. Virus-inclusive single-cell RNA sequencing reveals the molecular signature of progression to severe dengue. Proceedings of the National Academy of Sciences of the United States of America 2018.

7. Hottz ED, Oliveira MF, Nunes PC, et al. Dengue induces platelet activation, mitochondrial dysfunction and cell death through mechanisms that involve DC-SIGN and caspases. J Thromb Haemost 2013; 11:951–62.

8. Tassaneetrithep B, Burgess TH, Granelli-Piperno A, et al. DC-SIGN (CD209) mediates dengue virus infection of human dendritic cells. The Journal of experimental medicine 2003; 197:823–9.

9. Aguilar-Briseno JA, Upasani V, Ellen BMT, et al. TLR2 on blood monocytes senses dengue virus infection and its expression correlates with disease pathogenesis. Nat Commun 2020; 11:3177.

10. Yong YK, Tan HY, Jen SH, et al. Aberrant monocyte responses predict and characterize dengue virus infection in individuals with severe disease. Journal of translational medicine 2017; 15:121.

11. Hottz ED, Medeiros-de-Moraes IM, Vieira-de-Abreu A, et al. Platelet activation and apoptosis modulate monocyte inflammatory responses in dengue. J Immunol 2014; 193:1864– 72.

12. Malavige GN, Jeewandara C, Alles KM, et al. Suppression of virus specific immune responses by IL-10 in acute dengue infection. PLoS neglected tropical diseases 2013; 7:e2409.

13. Dayarathna S, Jeewandara C, Gomes L, et al. Similarities and differences between the ’cytokine storms’ in acute dengue and COVID-19. Sci Rep 2020; 10:19839.

14. Tsai TT, Chuang YJ, Lin YS, et al. Antibody-dependent enhancement infection facilitates dengue virus-regulated signaling of IL-10 production in monocytes. PLoS neglected tropical diseases 2014; 8:e3320.

15. Naranjo-Gomez JS, Castillo JA, Rojas M, et al. Different phenotypes of non-classical monocytes associated with systemic inflammation, endothelial alteration and hepatic compromise in patients with dengue. Immunology 2019; 156:147–63.

16. Duyen HTL, Cerny D, Trung DT, et al. Skin dendritic cell and T cell activation associated with dengue shock syndrome. Sci Rep 2017; 7:14224.

17. Lertjuthaporn S, Keawvichit R, Polsrila K, et al. Kinetic Changes of Peripheral Blood Monocyte Subsets and Expression of Co-Stimulatory Molecules during Acute Dengue Virus Infection. Pathogens 2021; 10.

18. Maheshwari D, Saini K, Singh P, et al. Contrasting behavior between the three human monocyte subsets in dengue pathophysiology. iScience 2022; 25:104384.

19. Durbin AP, Vargas MJ, Wanionek K, et al. Phenotyping of peripheral blood mononuclear cells during acute dengue illness demonstrates infection and increased activation of monocytes in severe cases compared to classic dengue fever. Virology 2008; 376:429–35.

20. WHO. Comprehensive guidelines for prevention and control of dengue fever and dengue haemorrhagic fever. SEARO Technical Publication Series. Vol. 60. SEARO, New Delhi, India: World Health Organization, 2011.

21. Obesity: preventing and managing the global epidemic. Report of a WHO consultation. World Health Organ Tech Rep Ser 2000; 894:i–xii, 1–253.

22. Amici SA, Young NA, Narvaez-Miranda J, et al. CD38 Is Robustly Induced in Human Macrophages and Monocytes in Inflammatory Conditions. Frontiers in immunology 2018; 9:1593.

23. Fleischer J, Soeth E, Reiling N, Grage-Griebenow E, Flad HD, Ernst M. Differential expression and function of CD80 (B7-1) and CD86 (B7-2) on human peripheral blood monocytes. Immunology 1996; 89:592–8.

24. Monneret G, Lafon T, Gossez M, et al. Monocyte HLA-DR expression in septic shock patients: insights from a 20-year real-world cohort of 1023 cases. Intensive care medicine 2025; 51:1820–32.

25. Chen Y, Zhang J, Cui W, Silverstein RL. CD36, a signaling receptor and fatty acid transporter that regulates immune cell metabolism and fate. The Journal of experimental medicine 2022; 219.

26. Zamora C, Canto E, Nieto JC, Angels Ortiz M, Juarez C, Vidal S. Functional consequences of CD36 downregulation by TLR signals. Cytokine 2012; 60:257–65.

27. Kuruppu H, Karunananda M, Jeewandara C, et al. Oxidative Stress Induced Liver Damage in Dengue Is Exacerbated in Those With Obesity. Open Forum Infect Dis 2025; 12:ofaf322.

28. Vuong NL, Lam PK, Ming DKY, et al. Combination of inflammatory and vascular markers in the febrile phase of dengue is associated with more severe outcomes. Elife 2021; 10.

29. Kuruppu H, De Silva M, Dissanayake C, et al. Functionality and phenotype of T cells in patients with varying severity of acute dengue and metabolic status. medRxiv 2025.

30. Gregorova M, Santopaolo M, Garner LC, et al. Early NK-cell and T-cell dysfunction marks progression to severe dengue in patients with obesity and healthy weight. Nat Commun 2025; 16:5569.

31. Dash MK, Samal S, Rout S, et al. Association of intermediate monocytes with dengue severity among the pediatric population of Odisha, India. Sci Rep 2025; 15:37116.

32. van de Weg CA, Pannuti CS, de Araujo ES, et al. Microbial translocation is associated with extensive immune activation in dengue virus infected patients with severe disease. PLoS neglected tropical diseases 2013; 7:e2236.

33. Wang WH, Lin CY, Chang K, et al. A clinical and epidemiological survey of the largest dengue outbreak in Southern Taiwan in 2015. Int J Infect Dis 2019; 88:88–99.

34. Chen PM, Katsuyama E, Satyam A, et al. CD38 reduces mitochondrial fitness and cytotoxic T cell response against viral infection in lupus patients by suppressing mitophagy. Sci Adv 2022; 8:eabo4271.

35. Heaton NS, Randall G. Dengue virus-induced autophagy regulates lipid metabolism. Cell host & microbe 2010; 8:422–32.

36. Metz P, Chiramel A, Chatel-Chaix L, et al. Dengue Virus Inhibition of Autophagic Flux and Dependency of Viral Replication on Proteasomal Degradation of the Autophagy Receptor p62. Journal of virology 2015; 89:8026–41.

37. Wijeratne DT, Fernando S, Gomes L, et al. Quantification of dengue virus specific T cell responses and correlation with viral load and clinical disease severity in acute dengue infection. PLoS neglected tropical diseases 2018; 12:e0006540.

38. Modhiran N, Watterson D, Muller DA, et al. Dengue virus NS1 protein activates cells via Toll-like receptor 4 and disrupts endothelial cell monolayer integrity. Science translational medicine 2015; 7:304ra142.

39. Thylur RP, Wu X, Gowda NM, et al. CD36 receptor regulates malaria-induced immune responses primarily at early blood stage infection contributing to parasitemia control and resistance to mortality. The Journal of biological chemistry 2017; 292:9394–408.

40. Olivetta E, Tirelli V, Chiozzini C, et al. HIV-1 Nef impairs key functional activities in human macrophages through CD36 downregulation. PloS one 2014; 9:e93699.

41. Rahimi N. C-type Lectin CD209L/L-SIGN and CD209/DC-SIGN: Cell Adhesion Molecules Turned to Pathogen Recognition Receptors. Biology (Basel) 2020; 10.

42. Sakuntabhai A, Turbpaiboon C, Casademont I, et al. A variant in the CD209 promoter is associated with severity of dengue disease. Nature genetics 2005; 37:507–13.

43. Cai G, Du M, Bosse Y, et al. SARS-CoV-2 Impairs Dendritic Cells and Regulates DC-SIGN Gene Expression in Tissues. International journal of molecular sciences 2021; 22.

44. Gopal R, Marinelli MA, Rago F, et al. CD209d/e promotes inflammation and lung injury during influenza virus infection. Immunohorizons 2025; 9.

45. Hsieh TH, Tsai TT, Chen CL, et al. Senescence in Monocytes Facilitates Dengue Virus Infection by Increasing Infectivity. Front Cell Infect Microbiol 2020; 10:375.

46. Patro ARK, Mohanty S, Prusty BK, et al. Cytokine Signature Associated with Disease Severity in Dengue. Viruses 2019; 11.

47. Tian N, Zheng JX, Guo ZY, et al. Dengue Incidence Trends and Its Burden in Major Endemic Regions from 1990 to 2019. Trop Med Infect Dis 2022; 7.

48. Azeredo EL, Neves-Souza PC, Alvarenga AR, et al. Differential regulation of toll-like receptor-2, toll-like receptor-4, CD16 and human leucocyte antigen-DR on peripheral blood monocytes during mild and severe dengue fever. Immunology 2010; 130:202–16.

