## Supplementary methods for "An early activated CD38^high^CD209^high^CD36^low^ monocyte phenotype is associated with progression to severe dengue"

| Antibody | Fluorochrome | Clone | Vendor |
|---|---|---|---|
| CD14 | FITC | M5E2 | BD Pharmingen |
| CD16 | Pacific Blue | 3G8 | BioLegend, USA |
| CD209 | PEcy7 | 9E9A8 | BioLegend, USA |
| CD86 | PE | W17233E | BioLegend, USA |
| Viability dye | BV510 | - | BioLegend, USA |
| HLA-DR | APC | LN3 | BioLegend, USA |
| CD38 | AF700 | HB-7 | BioLegend, USA |
| CD36 | APC-Cy7 | 5-271 | BioLegend, USA |
| CD3 | BV605 | OKT3 | BioLegend, USA |
| CD19 | BV605 | HIB19 | BioLegend, USA |
| CD56 | BV605 | HCD56 | BioLegend, USA |
| CD15 | BV605 | W6D3 | BioLegend, USA |

Table 1:

**Hi-dimensional data analysis**

*Data Preprocessing and Transformation*

Raw Flow Cytometry Standard (.fcs) files of samples of patients with dengue infection (DF, n=33 and DHF, n=21) and samples of healthy individuals (n=20) were analyzed and concatenated as two separate Flow Cytometry Standard (.fcs) files (Total patient concat. and Healthy group concat.). Those concatenated Flow Cytometry Standard (.fcs) files were subjected to Down sampling and batch effect correction using Down sample plugin and Cycombine plugin in FlowJo software (version10). Preprocessed concatenated Flow Cytometry Standard (.fcs) files were imported into biomeStat: Version 1.0. (Bioinformatics Analysis Platform) and parsed using the Flowjo library in Python. To account for the log-normal distribution of fluorescence data and handle negative values introduced during compensation, marker intensities were transformed using an inverse hyperbolic sine (arcsinh) function with a cofactor of 150, a standard setting for flow cytometry data.

*Dimensionality Reduction (UMAP)*

To visualize the high-dimensional structure of the data in two dimensions, we applied the Uniform Manifold Approximation and Projection (UMAP) algorithm (McInnes et al., 2018). The analysis was performed on the transformed expression data of seven key markers: CD14 (FITC), CD16 (Pacific Blue), HLA-DR (APC), CD36 (APC Cy7), CD86 (PE), CD209 (PE Cy7), and CD38 (Alexa Fluor700). UMAP parameters were set to n_neighbors=15 to preserve local data structure and min_dist=0.1 to control the tightness of embedding points.

Resulting UMAP of the patient group was overlaid with **Severity Group** metadata, visualizing samples of Dengue Fever (DF) (green) and samples of Dengue Hemorrhagic Fever (DHF) (red). Additionally, resulted UMAPs were overlaid with **Obesity Group** metadata to visualize samples of Lean (yellow) and samples of Obese (purple) across the UMAP landscape of both total patient group and the healthy control group.

*Clustering and Population Identification (FlowSOM)*

To identify distinct monocyte subsets, clustering was performed specifically on the CD14 (FITC) and CD16 (Pacific blue) channels. A K-Means clustering algorithm (k=4) was applied to the transformed data, functionally determining the "meta-clusters" akin to the FlowSOM workflow (Van Gassen et al., 2015) in biomeStat: Version 1.0. (Bioinformatics Analysis Platform) to partition the events into three distinct populations based on their expression profiles. Clusters were annotated based on the biological definitions of monocyte subsets:

1. **Classical Monocytes**: High CD14 expression, low CD16 expression (Aquamarine Green).
2. **Non-classical Monocytes**: High CD16 expression, low CD14 expression (Blue).
3. **Intermediate Monocytes**: Intermediate CD14 and CD16 expression (Red).

The three clusters were subsequently overlayed onto the original UMAP of both total patient group and the healthy control group to visualize their spatial distribution.

**References**


1. Caleyachetty R., Barber T.M., Mohammed N.I., Cappuccio F.P., Hardy R., Mathur R., Banerjee A. and Gill P. Ethnicity-specific BMI cutoffs for obesity based on type 2 diabetes risk in England: a population-based cohort study. Lancet Diabetes Endocrinol 9(7):419-426, 2021.

2. Marimuthu R, Francis H, Dervish S, Li SCH, Medbury H, Williams H. Characterization of human monocyte subsets by whole blood flow cytometry analysis. *J Vis Exp*. 2018;(140):e57941. doi:10.3791/57941.
3. Manichaikul A, Rich SS, Perry H, Yeboah J, Law M, Davis M, et al. *Supplementary Methods: Monocyte subsets*. NIH Manuscript Submission; NIHMS943140.
4. **UMAP:** McInnes, L., Healy, J., & Melville, J. (2018). Umap: Uniform manifold approximation and projection for dimension reduction. *arXiv preprint arXiv:1802.03426*.
5. **FlowSOM:** Van Gassen, S., Callebaut, B., Van Helden, M. J., ... & Saeys, Y. (2015). FlowSOM: Using self-organizing maps for visualization and interpretation of cytometry data. *Cytometry Part A*, 87(7), 636-645.
