## Supplementary data for "An early activated CD38^high^CD209^high^CD36^low^ monocyte phenotype is associated with progression to severe dengue"

| | DF<br><br>N=33 (%) | DHF<br><br>N=21 (%) |
|---|---|---|
| **Gender**<br><br>Male<br><br>Female | <br><br>17 (51.5%)<br><br>16 (48.5%) | <br><br>13 (61.9%)<br><br>08 (38.1%) |
| **Age:** Median (IQR) | 26 (20 – 41) | 28 (22 – 43) |
| **Days since symptoms:** Median (IQR) | 4 (3 – 4) | 4 (4 – 5) |
| **Fever** | 33 (100%) | 21(100%) |
| **Abdominal pain** | 8 (24.24%) | 10 (47.6%) |
| **Vomiting** | 10 (30.3%) | 7 (33.3%) |
| **Pericholecystic fluid** | 0 (0%) | 21(100%) |
| **Ascites** | 0 (0%) | 17 (80.95%) |
| **Pleural effusion** | 0 (0%) | 2 (9.52%) |
| **Bleeding manifestations** | 2 (6.1%) | 2 (9.5%) |
| **Leucopenia** <4,000 cells (μ L) | 25 (75.75%) | 16 (76.2%) |
| **Platelets count on admission** cells/mm$^3$:<br><br>Median (IQR) | 140 (120 – 164) | 102 (64 – 141) |

| | | |
|---|---|---|
| **Lowest Platelets count cells/mm$^3$**: Median (IQR) | 98 (79 – 121) | 36 (14 – 43) |
| **AST IU/L:** Median (IQR) | 62 (30 – 105) | 93 (58 – 134) |
| **ALT IU/L:** Median (IQR) | 67 (31 - 116) | 70 (46.5 – 111) |
| **CRP** ng/L: Median (IQR) | 11.2 (6.8 – 21.5) | 20.5 (8.7 – 32.7) |
| **NS1 rapid test +** | 19/33 (57.6%) | 14/21 (66.7%) |

**Table 1: Clinical and demographic characteristics of patients**
