## supplementary figures for "An early activated CD38^high^CD209^high^CD36^low^ monocyte phenotype is associated with progression to severe dengue"

Figure S1

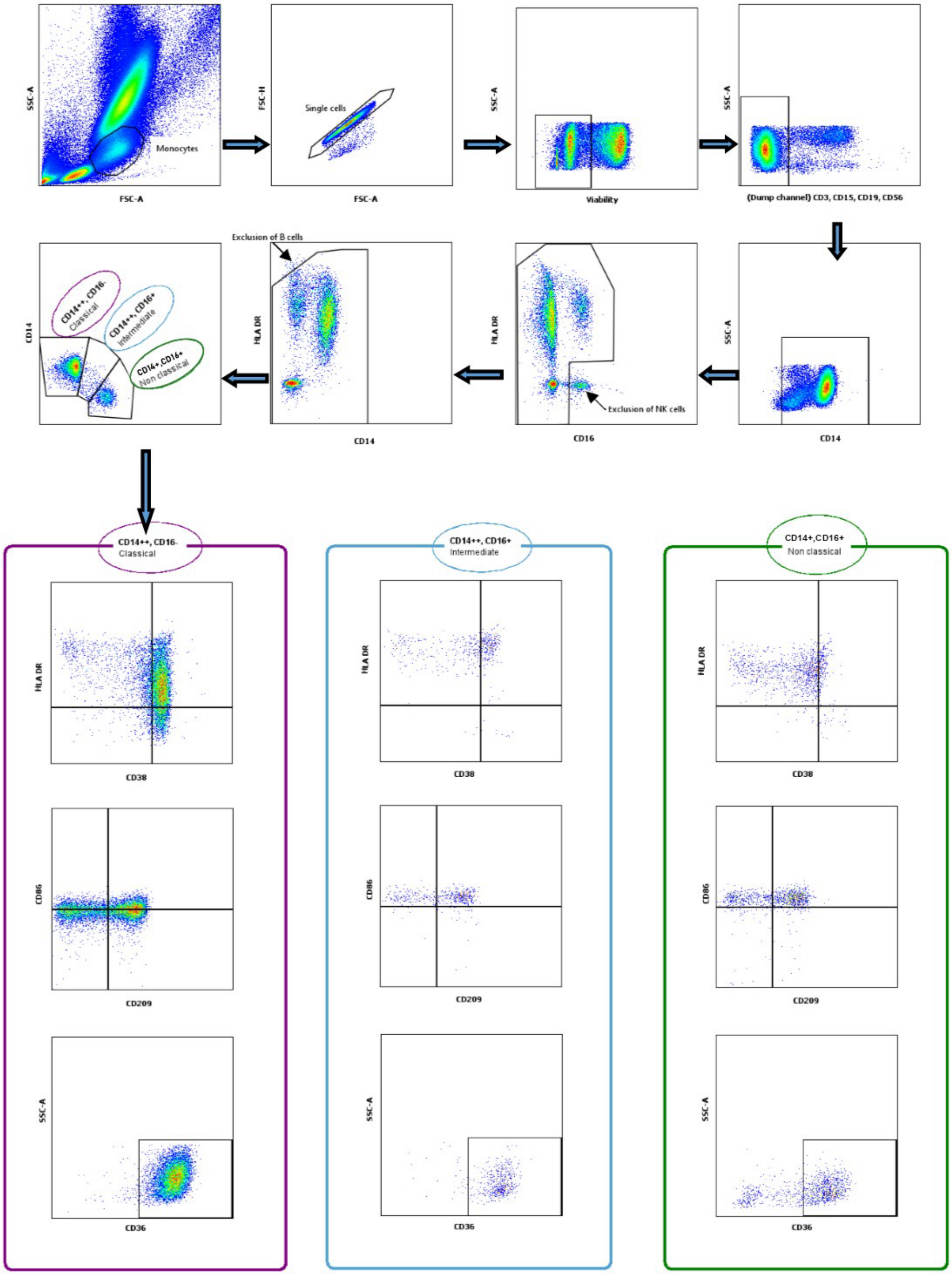

SSC-A
FSC-A
Monocytes
FSC-H
Single cells
Viability
(Dump channel) CD3, CD15, CD19, CD56
CD14
HLA DR
CD16
Exclusion of NK cells
Exclusion of B cells
CD14++, CD16-
Classical
CD14++, CD16+
Intermediate
CD14+,CD16+
Non classical
CD38
CD86
CD209
CD36

Figure S2

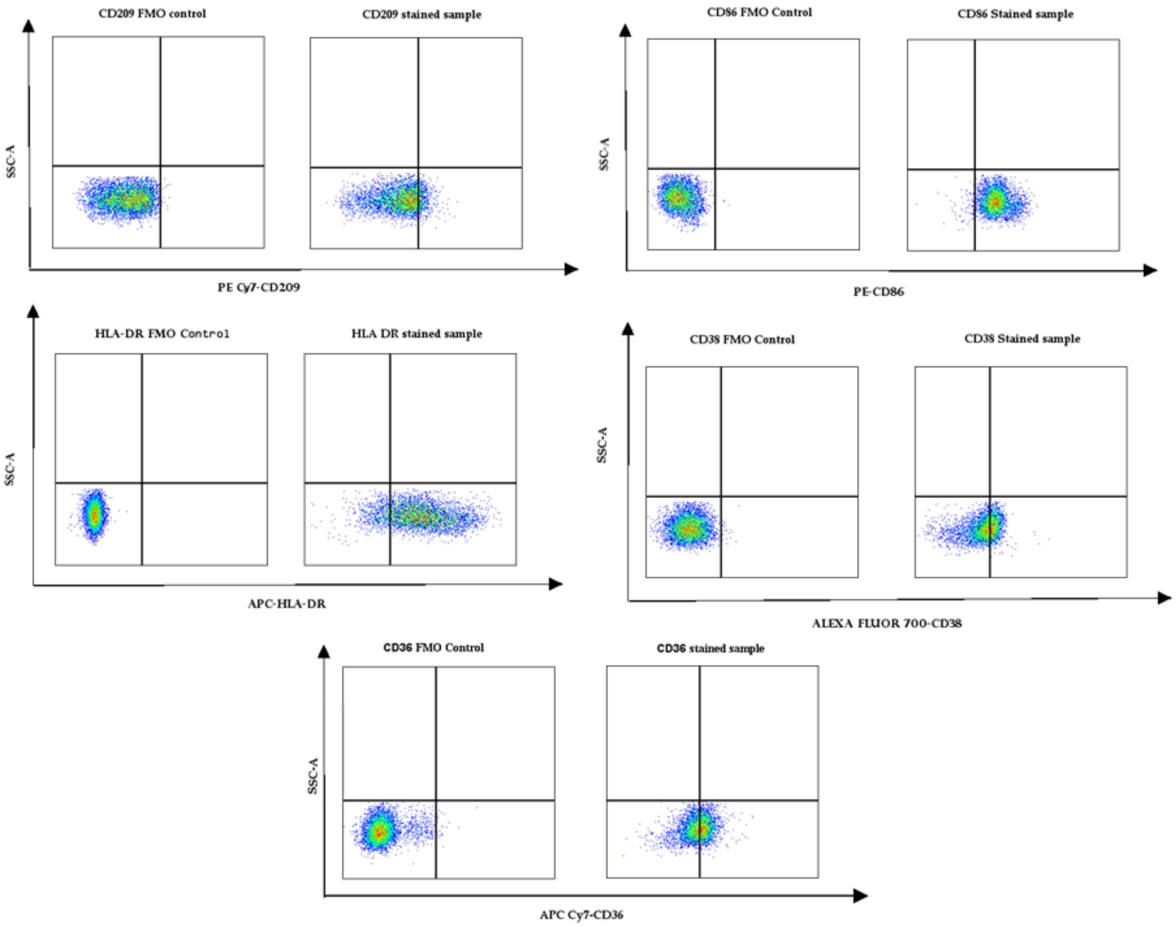
CD209 FMO control
CD209 stained sample
SSC-A
PE Cy7-CD209
CD86 FMO Control
CD86 Stained sample
SSC-A
PE-CD86
HLA-DR FMO Control
HLA DR stained sample
SSC-A
APC-HLA-DR
CD38 FMO Control
CD38 Stained sample
SSC-A
ALEXA FLUOR 700-CD38
CD36 FMO Control
CD36 stained sample
SSC-A
APC Cy7-CD36

Figure S3

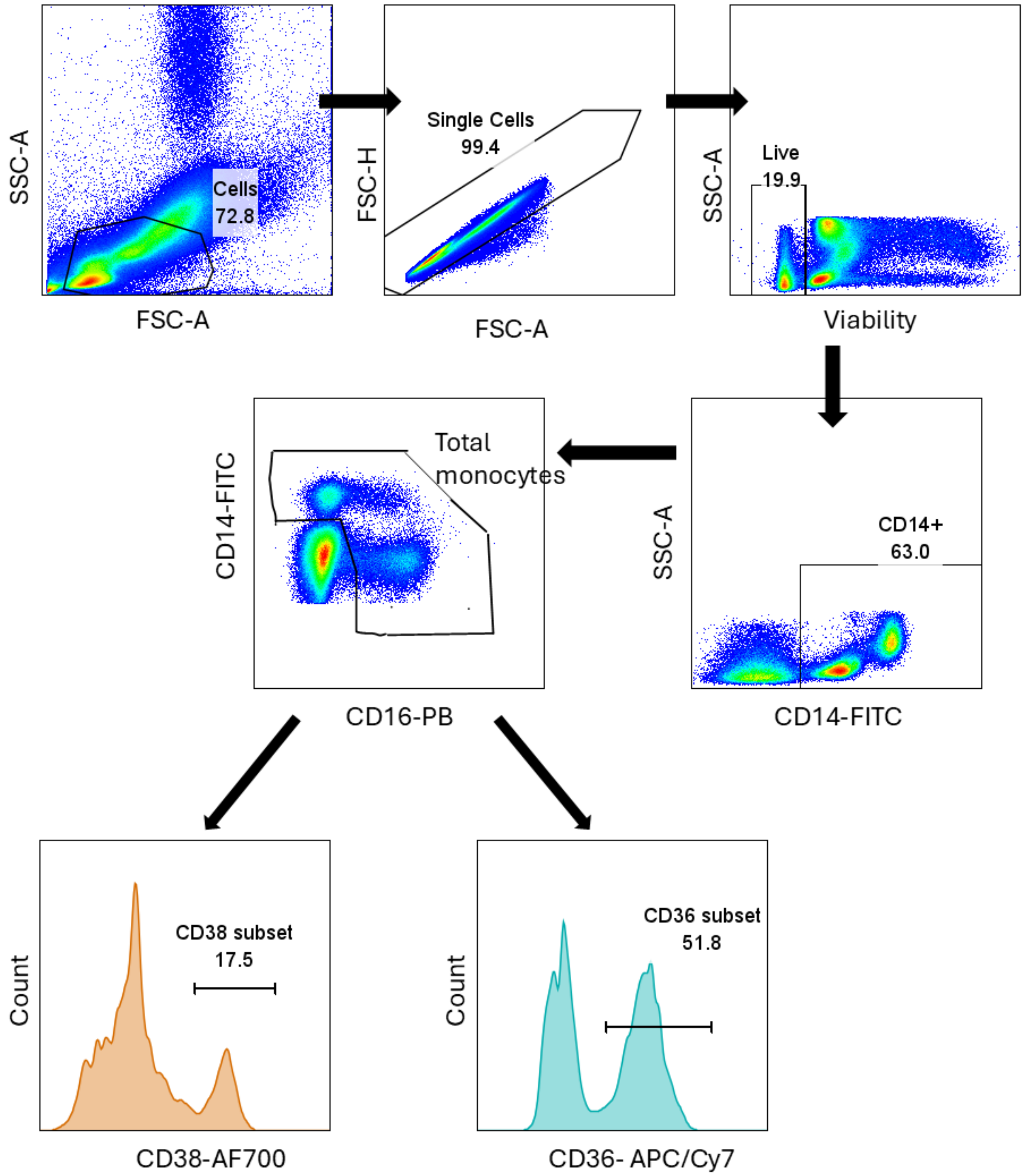

SSC-A
Cells
72.8
FSC-A
FSC-H
Single Cells
99.4
FSC-A
SSC-A
Live
19.9
Viability
SSC-A
CD14+
63.0
CD14-FITC
CD14-FITC
Total
monocytes
CD16-PB
Count
CD38 subset
17.5
CD38-AF700
Count
CD36 subset
51.8
CD36- APC/Cy7
